# Novel Large Language Model-Based Detection of Echocardiographic Markers of Right Ventricular Dysfunction

**DOI:** 10.64898/2026.08.26.26361456

**Authors:** Leela Ekambarapu, Akshay Pendyal, Anthony Lin, Mahmoud Alwakeel, Arun Rajaratnam

**Affiliations:** Duke University School of Medicine, Department of Medicine; Durham, NC; Duke University School of Medicine, Department of Medicine, Division of Cardiology; Durham, NC; Duke University School of Medicine, Department of Medicine, Division of General Internal Medicine; Durham, NC

## Abstract

**Background:** Unstructured biomedical data, such as echocardiography reports, are rich in information but time consuming to analyze at scale. Rule-based, regular expression–driven terminology mapping can only extract individual variables while large language models (LLMs) offer scalable and clinically meaningful interpretations of heterogeneous disease processes. Right ventricular dysfunction (RVD) is an example of a multifactorial disease state in which key structural and physiologic features are captured both narratively and in structured fields, making it an ideal test case for evaluating whether LLMs can recover complex phenotypes that rules-based methods routinely miss.

**Purpose:** To compare an LLM-based extraction method to a conventional rules-based schema for identifying and phenotyping echocardiographic features associated with RVD in a large TTE dataset.

**Methods:** MIMIC-III NOTE2NUM echocardiography reports (n = 45,794) were analyzed using GPT-4o– based LLM extraction deployed within a secure health system enclave and were benchmarked against echocardiographic measurements defined in the MIMIC-III dictionary schema. In MIMIC-III, PH was recorded qualitatively (mild/moderate/severe) based on tricuspid regurgitant (TR) jet velocity and then recoded as present vs. absent. LLM based extraction defined RVD as (1) RV structural abnormality (≥1 of hypertrophy, dilation, or wall hypo-/akinesis) or (2) RV pressure/volume overload (≥2 of the following: estimated right atrial pressure > 8 mmHg, TR jet velocity > 2.8 m/s, fractional area change < 35%, tricuspid annular planar systolic excursion < 17 mm, S′ < 9.5 cm/s, or E/e′ > 14), with PH defined as estimated pulmonary artery systolic pressure > 35 mmHg or qualitative documentation of PH.

**Results:** LLM extraction identified PH in 15,394 (33.6%), RV pressure/volume overload in 14,449 (31.6%), and RV structural abnormalities in 11,955 (26.1%). Co-occurrence was common: overload + structural changes in 9,380 (20.5%), overload + PH in 9,756 (21.3%), structural changes + PH in 6,183 (13.5%), and all three in 5,620 (12.3%). Using the MIMIC-III dictionary schema, PH prevalence was similar (15,371; 33.6%), but RV overload fields were captured less often (pressure overload 1,357 [3.0%], volume overload 1,128 [2.5%], pressure + volume overload 1,093 [2.4%]; any overload field 3,578 [7.8%]), and RV pressure/volume overload with PH was identified in only 731 (1.6%).

**Conclusions:** LLM-based extraction outperforms rules-based schemas for identifying complex disease states not defined by any single variable. By synthesizing multifactorial signals, LLMs can phenotype RVD with higher fidelity and support population-level assessment. Further validation using multimodality imaging, invasive hemodynamics, and clinical outcome data is needed.

## Introduction

Modern healthcare systems generate vast volumes of unstructured clinical text, creating a growing need for scalable tools that can transform narrative documentation into structured, analyzable data. Natural language processing (NLP) has long been used in medicine for automated extraction of data from various unstructured texts including clinical notes, surgical reports, and radiology results. However, NLP relies on dictionary-driven schemas and identification of pre-defined labels which often require manual curation, rigid rule construction, and extensive maintenance to keep pace with evolving clinical language.^4^ These features make NLP ill-suited for capturing multifactorial disease processes.

In contrast, large language models (LLMs) have emerged within the last decade as a robust and scalable approach to extracting clinically meaningful constructs. Rather than relying on fixed dictionaries, LLMs can integrate multiple interdependent descriptors and interpret text holistically.^15^ Early applications of LLMs in clinical imaging and text analyses demonstrate promising diagnostic and phenotyping capabilities, but systematic evaluation of their performance for complex, multifactorial disease extraction has been limited.^2,13^

Right ventricular dysfunction (RVD) is an example of a heterogenous disease process that is underrecognized but is a major contributor to adverse outcomes across a wide range of cardiopulmonary disease states. RVD is associated with poor functional status and confers a heightened risk of mortality irrespective of underlying etiology, highlighting its role as a distinct and clinically meaningful contributor to adverse outcomes. As conditions that disproportionately affect the RV, such as pulmonary hypertension (PH), are becoming increasingly common, earlier detection and subsequent management of RVD is critical.^9,12^

Evaluation of RV performance can be done with several complementary methods. Defined here as structural abnormalities accompanied by evidence of pressure and volume overload, RVD is most accurately characterized by invasive hemodynamic assessment via right heart catheterization. Multimodal imaging techniques serve as noninvasive avenues to assess RV function. Notably, transthoracic echocardiography (TTE) remains the initial imaging modality of choice to evaluate RV function given its widespread availability, portability, low cost, and lack of ionizing radiation exposure. However, depending on the underlying disease process of interest, TTE-derived measurements of RV function are variably predictive due to the chamber’s complex geometry and heterogenous remodeling response to pressure or volume stress.^5,8^ Nevertheless, TTE reports represent a large reservoir of unstructured textual data, containing narrative descriptions that often capture nuanced clinical impressions, qualitative assessments, and contextual details that are not reflected in structured fields.^4^

While LLMs have emerged as a promising tool for synthesizing complex information and identifying patterns, to our knowledge, there has been no direct comparison between LLMs and a conventional dictionary schema in identifying RVD from standard, unstructured TTE reports. In this study, we used ConceptExtract.AI, a proprietary, internally developed and validated LLM-based concept extraction platform, to operationalize structured phenotyping of RVD from free-text echocardiographic reports at scale, comparing its yield against a conventional rules-based schema. We further applied this approach to stratify RVD by the presence or absence of pulmonary hypertension (PH) and across a range of left ventricular ejection fraction (EF) categories, enabling phenotypic characterization across clinically meaningful subgroups.

## Methods

### Data

Echocardiography reports used for this study were obtained from the MIMIC-III (ECHO-NOTE2NUM) database. This is a large dataset (n = 45,794) of publicly available and deidentified echocardiogram reports conducted in various intensive care units at Beth Israel Deaconess Medical Center from 2001 to 2012.^11^ This study was exempted from IRB review.

### ConceptExtract.AI

In the MIMIC-III dictionary schema, PH was recorded qualitatively as mild, moderate, or severe based on tricuspid regurgitant (TR) jet velocity and then re-coded as a binary indicator (present vs. absent). For right ventricle volume overload and dilation, the hierarchical numerical coding system was applied with the schema shown in Figure 2 as follows; -3 (not specifically mentioned, difficult to view), 0 (not overload/dilated), 1 (overload/dilated). Pressures were coded as -3 (specifically mentioned as indeterminate or difficult to view), 0 (normal), or 1 (abnormal). If there was no mention of function or dysfunction for a given concept, the field was left null.^11^ LLM-based extraction defined RVD as (1) RV structural abnormality (≥1 of hypertrophy, dilation, or hypo- or akinetic wall motion) or (2) RV pressure/volume overload (≥2 of estimated right atrial pressure (RAP) > 8 mmHg, TR jet velocity > 2.8 m/s, fractional area change (FAC) < 35%, tricuspid annular planar systolic excursion (TAPSE) < 17 mm, S′ < 9.5 cm/s, or E/e′ > 14).^14^ PH was defined as estimated pulmonary artery systolic pressure (ePASP) > 35 mmHg or qualitative documentation of PH (e.g. “The constellation of findings is concerning for pulmonary hypertension.”).

### Platform Description

ConceptExtract.AI is an internally developed, no-code LLM-based platform for structured concept extraction from unstructured clinical text, designed to be accessible to clinical researchers without programming expertise. The platform accepts a user-defined concept-definition layer in which each clinical concept is paired with a free-text operationalization and produces for every input document a structured prediction per concept comprising a categorical label drawn from a user-specified label space and a mandatory supporting rationale citing the source text. All concepts are extracted in a single batched LLM call per document, ensuring each concept is evaluated against the same input window. ConceptExtract.AI incorporates a multi-layered quality assurance architecture (Figure 1). In the primary extraction pass, mandatory rationale generation anchors every label to the source text. A second independent LLM then re-evaluates each prediction against the source document and concept definition, a dual-model concordance design in which concordant predictions proceed automatically while discordant cases are flagged for further review. Flagged records can be escalated to a third independent model for adjudication, providing an additional layer of automated oversight before any residual disagreement is routed for targeted human review. This tiered concordance approach mirrors established frameworks for LLM-based clinical abstraction quality assurance, in which dual-model agreement has been shown to achieve positive predictive value ≥0.95 and negative predictive value ≥0.98 against a human gold standard.^1^

**Figure 1.**
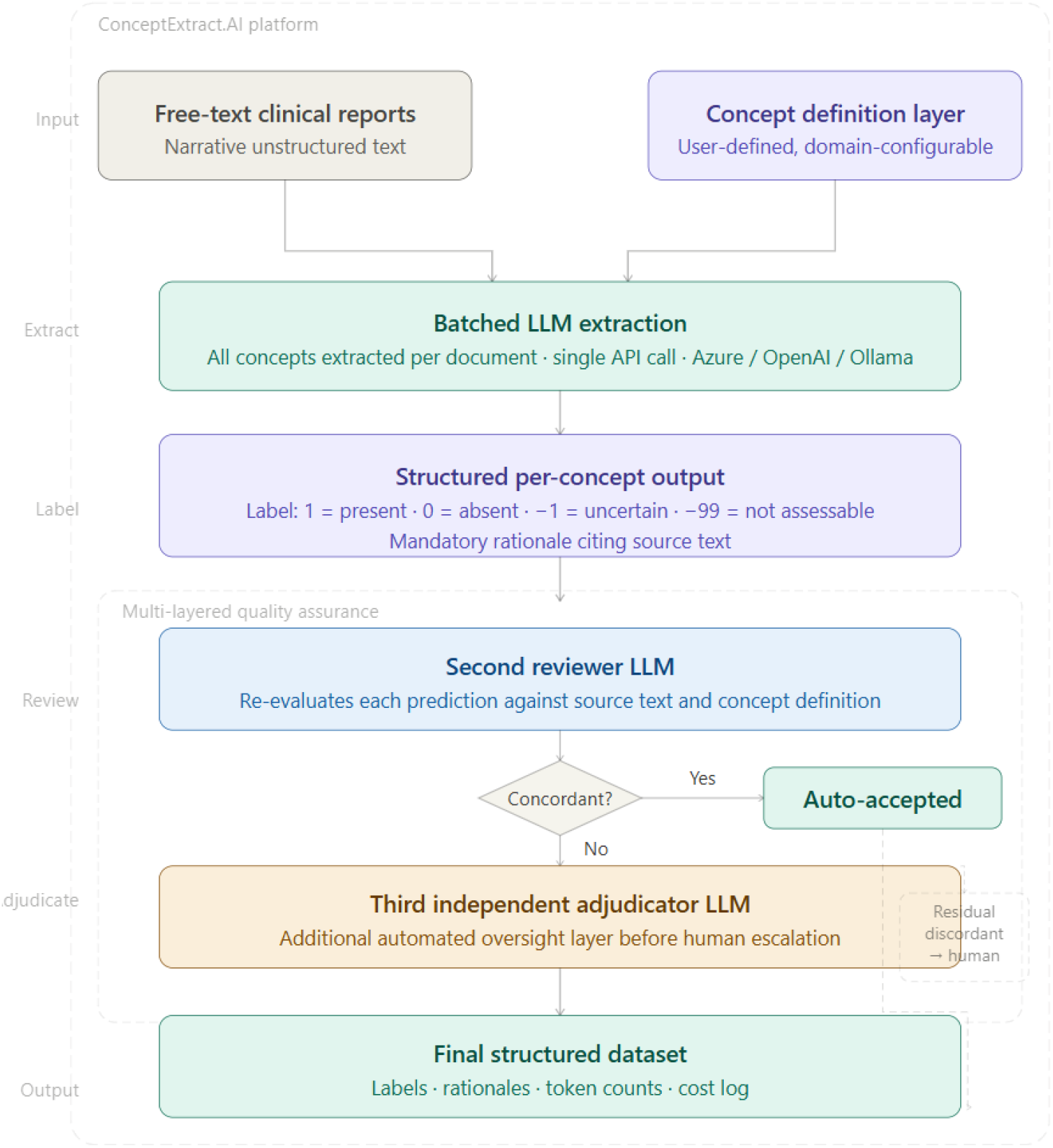
Schematic Illustration of ConceptExtract.AI Development Process. This figure illustrates how.AI consists of three sequential LLM-based extractions, multi-layered quality assurance, and generation of a validated structured dataset from free-text clinical reports.

**Figure 2.**
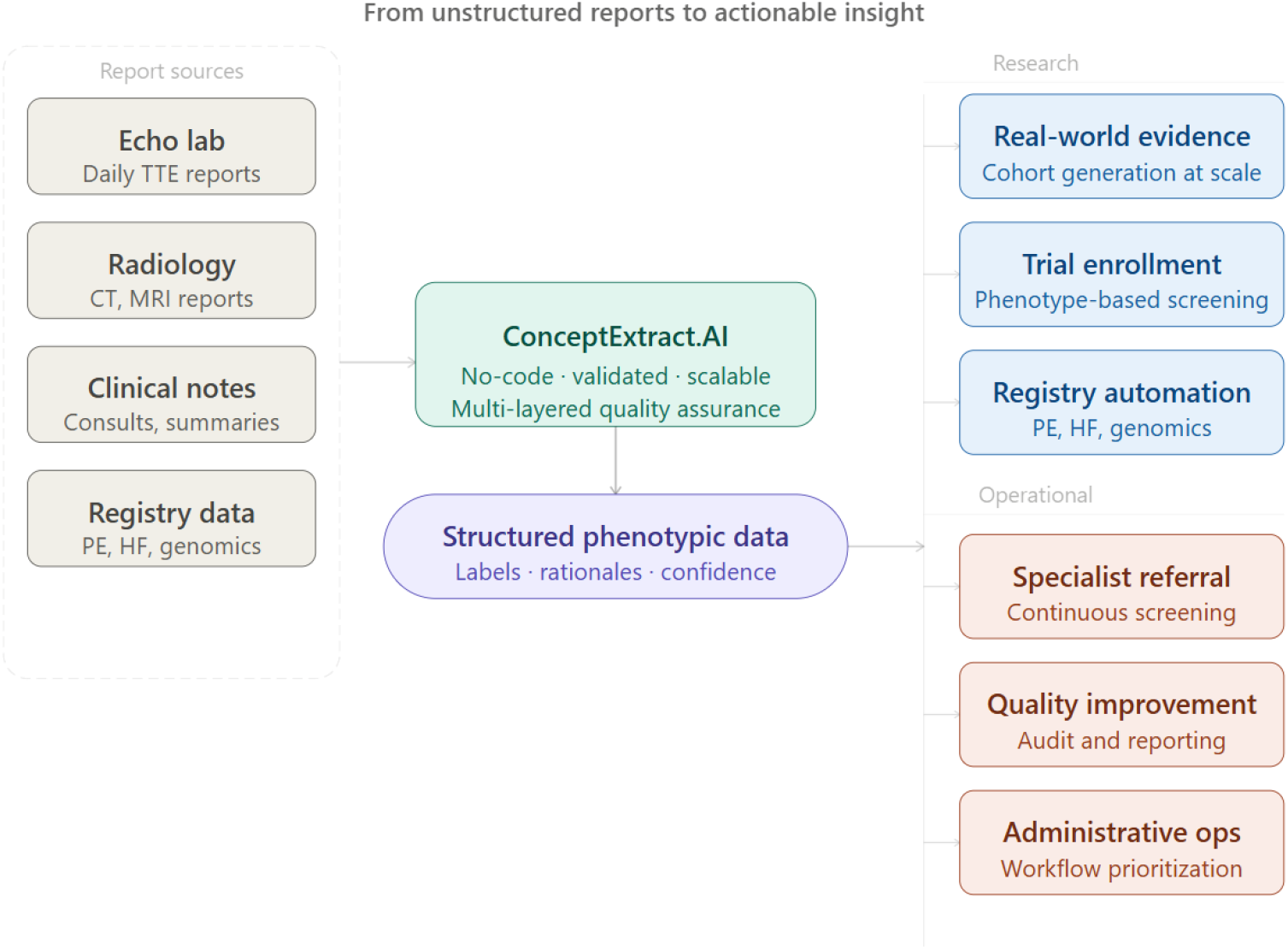
ConceptExtract.AI Pipeline from Unstructured Text to Structured Outputs. High-level overview showing how diverse unstructured clinical reports are processed through ConceptExtract.AI to generate validated structured phenotypic data that support downstream research and operational applications.

The platform has been internally validated against human-annotated datasets across multiple clinical domains, including cardiovascular medicine, pulmonology, psychiatry, neuroscience, and obstetrics and gynecology; it is currently deployed in active research programs in genomics and additional clinical specialties (Figure 2). The platform supports OpenAI, Azure OpenAI, and locally hosted model deployments, enabling use across environments with different data-governance requirements. Deterministic checkpointing and per-record cost logging support reliable execution at population scale.

## Results

### Baseline Extraction with Dictionary Schema

Using a rules-based/dictionary schema, of the 45,794 reports analyzed, PH was present in 15,371 (33.6%). RV pressure overload was captured in 1,357 (3.0%), while RV volume overload was present in 1,128 (2.5%). Co-occurrence (pressure and volume overload) was present in 1,093 (2.4%) while the presence of any overload field was 3,578 (7.8%). The combination of RV pressure/volume overload and PH was identified in 731 (1.6%).

### Comparison of LLM Based Extraction

LLM extraction identified PH in 15,394 (33.6%), similar in performance to the dictionary schema. In contrast, RV pressure/volume overload was present in 14,449 (31.6%). RV structural abnormalities were present in 11,955 (26.1%). Co-occurrence of features of RVD (structural change plus pressure/volume overload) and PH was common: RV pressure/volume overload plus structural changes in 9,380 (20.5%), RV pressure/volume overload plus PH in 9,756 (21.3%), RV structural changes plus PH in 6,183 (13.5%), and all three in 5,620 (12.3%) (Figure 3).

**Figure 3.**
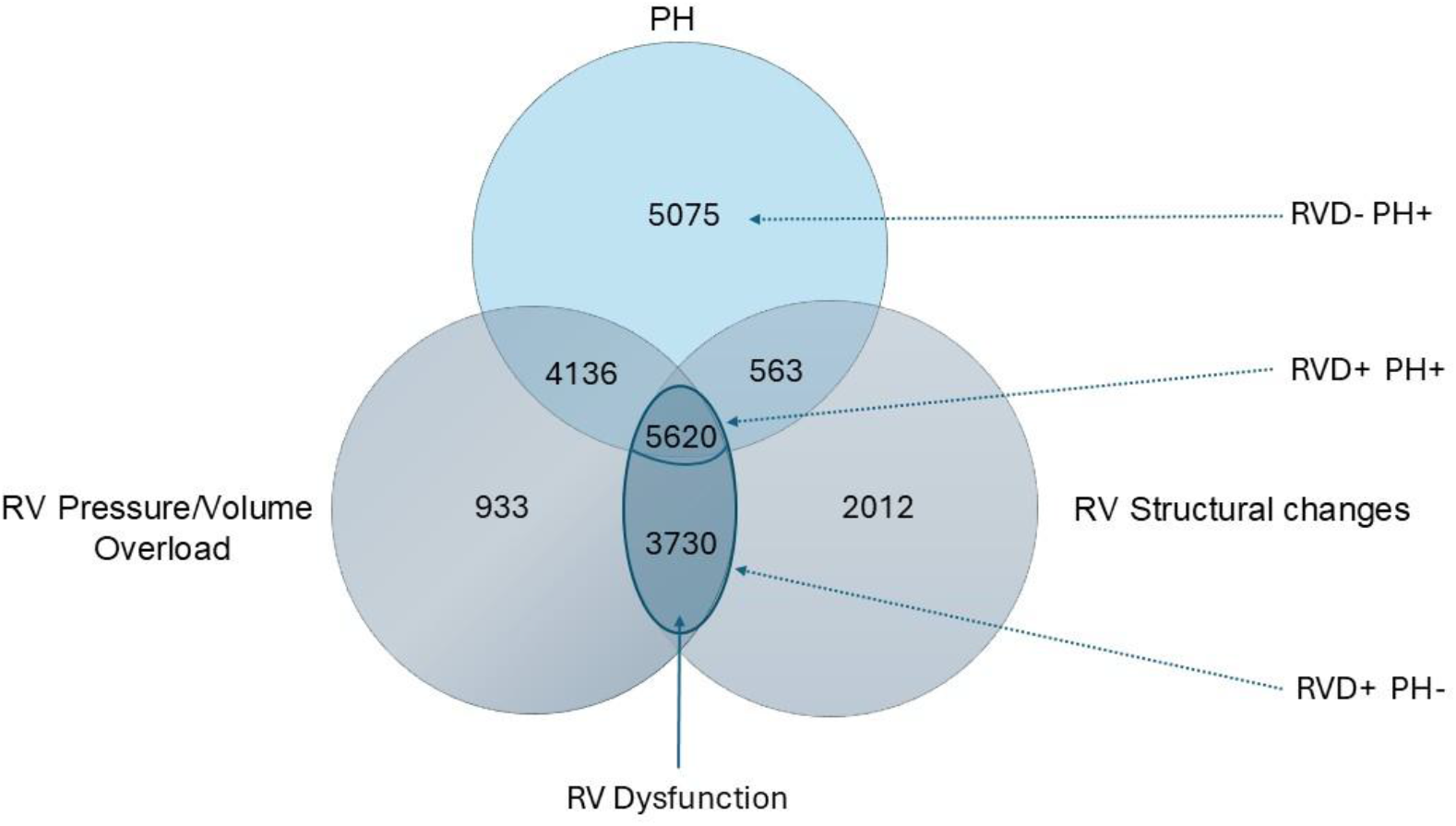
RV Phenotypes on Echocardiography Based on LLM definitions. Counts in the Venn diagram indicate the number of studies meeting criteria for each phenotype and their co-occurrence. The central shaded region highlights cooccurrence of RVD and PH, demonstrating the multifactorial nature of RVD. Abbreviations: RV: Right Ventricle; RVD; Right Ventricular Dysfunction defined as ≥2 pressure, area, based, or doppler-based physiological changes and the presence of structural changes of the RV; PH: Pulmonary Hypertension, defined as estimated pulmonary artery pressure (ePASP) >35mmHg or qualitative reporting of PH (mild/moderate/severe).

### Stratification by Ejection Fraction

The presence of RVD and PH was further stratified by left ventricular EF. Notably, 27% of all echo reports in the MIMIC-III database were missing EF quantification due to technical limitations or having been performed as focused studies. In those with a normal EF (>50%; n = 27,082), abnormal RV findings were common with any abnormal RV measure (structural change or pressure overload/volume overload) documented in 8,504 (31%) and RV pressure/volume overload documented specifically in 7,041 (26%), while RVD was present in 3,846 (14%). Co-occurrence of RVD and PH was present in 2,573 (10%). RVD without PH was present in 1,273 (4.7%) while PH without RVD was present in 3,602 (13.3%) (Table 1).

**Table 1.** Distribution of Right Ventricular Abnormalities and Pulmonary Hypertension Stratified by Left Ventricular Ejection Fraction. Percentages represent the proportion of patients within each EF group meeting criteria for right ventricular abnormalities or pulmonary hypertension; corresponding sample sizes are shown in parentheses. Abbreviations: PH: Pulmonary Hypertension, RVD: Right Ventricular Dysfunction, P/V overload: Pressure/Volume Overload; Str: Structural.

| EF | RVD-<br>PH+ | RVD+<br>PH- | RVD+<br>PH+ | RVD<br>(total) | RV P/V<br>overload | RV Str<br>Change | Any RV<br>abnormal<br>measure |
| --- | --- | --- | --- | --- | --- | --- | --- |
| >50%<br>(n=27,082) | 13.3%<br>(n=3,602) | 4.7%<br>(n=1,273) | 9.5%<br>(n=2,573) | 14.2%<br>(n=3,846) | 26.0%<br>(n=7,041) | 19.6%<br>(n=5,308) | 31.4%<br>(n=8,504) |
| 41-49%<br>(n=1,189) | 7.5%<br>(n=89) | 12.0%<br>(n=143) | 13.7%<br>(n=163) | 25.7%<br>(n=306) | 34.2%<br>(n=407) | 34.8%<br>(n=414) | 43.2%<br>(n=514) |
| <40%<br>(n=5,126) | 8.1%<br>(n=415) | 19.2%<br>(n=984) | 26.9%<br>(n=1,379) | 46.1%<br>(n=2,363) | 56.1%<br>(n=2,876) | 25.3%<br>(n=1,297) | 36.8%<br>(n=1,886) |

Reduced EF was further divided into EF 41-49% (mildly reduced) and EF <40% (moderate to severely reduced). In those with mildly reduced EF (n = 1,189), any abnormal RV measure was present in 514 (43.2%) while RVD was present in 306 (25.7%). RVD without PH was present in 143 (12.0%) while PH without RVD was present in 89 (7.5%). RVD plus PH was present in 163 (13.7%). In those with moderate to severely reduced EF (n = 5,126), any abnormal RV measure was present in 1,186 (36.8%) while RVD was present in 2,363 (46.1%). RVD without PH was present in 984 (19.2%) while PH without RVD was present in 415 (8.1%). RVD plus PH was present in 1,379 (26.9%) (Table 1).

In those classified as heart failure with preserved ejection fraction (HFpEF) based on International Classification of Disease, Ninth Revision (ICD-9) coding (n = 10,062), any abnormal RV measure was present in 6,746 (67%) with RVD present in 2,926 (29.1%). RVD without PH was present in 696 (6.9%), PH without RVD was present in 1,456 (14.5%) and co-occurrence of RVD and PH was present in 2,230 (22.2%).

### Prevalence of RVD and PH Over Time

In patients with HFpEF with more than three TTEs in the MIMIC-III database, initial and final TTEs were compared to determine the change in prevalence of RVD and PH. The presence of PH without RVD decreased 2.11% from initial to final TTE (p = 0.17). RVD without PH increased 2.30% from initial to final TTE (p < 0.0001) and co-occurrence of RVD and PH increased 7.29% (p < 0.0001) (Figure 4).

**Figure 4.**
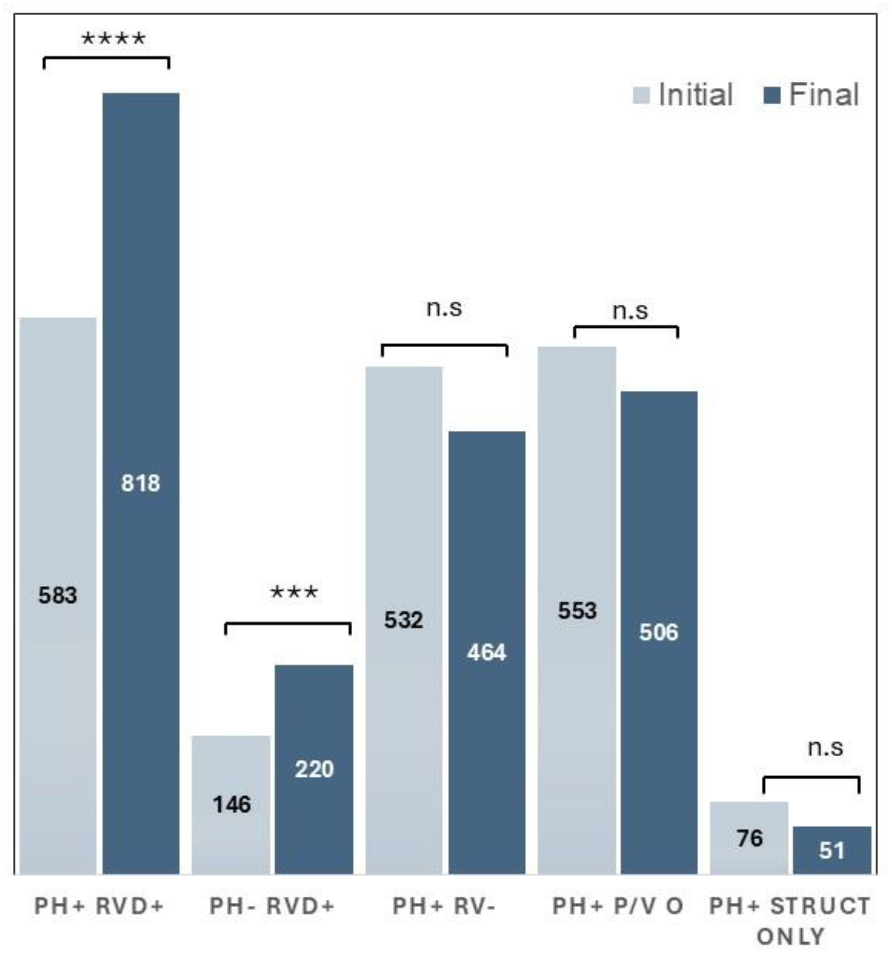
Proportion of RVD and PH Subtypes in Initial versus Final Echocardiograms in HFpEF. Statistically significant increase in the number of patients with RVD with and without PH. Abbreviations: PH: Pulmonary Hypertension, RVD: Right Ventricular Dysfunction, P/V O: Pressure/Volume Overload.

## Discussion

Findings from this study highlight the enhanced ability of LLMs, compared to traditional rules-based schemas, to identify heterogenous and multifactorial disease states such as RVD on a large-scale basis. This adds to the growing evidence that AI methods can be employed to extract clinically meaningful data from unstructured sources.^2^ To our knowledge, this is the first study using LLM-based methods to evaluate RVD holistically as a combination of both structural abnormalities and RV changes associated with pressure and volume overload seen on echocardiography.

In our study, the patterns extracted by the LLM elicit a far broader and more nuanced representation of RVD than what is ordinarily achievable with rules-based NLP. While elevated pulmonary pressures and increased RV afterload remain important contributors to RV dysfunction, our findings demonstrate that RVD frequently occurs even in the absence of echocardiographic evidence or qualitative documentation of pulmonary hypertension. These results suggest that macrostructural changes in RV function are likely to occur via mechanisms not dependent on pulmonary vascular loading.^7,9^ The ability of the LLM to detect these patterns, despite their often variable documentation, illustrates the advantage of contextual interpretation over rules-based identification.

Our study also demonstrates the potential utility of an LLM-based approach in identifying RVD with concurrent HF, across a range of left ventricular EF. Prior work has demonstrated that RVD is common in both HFrEF and HFpEF, and carries important prognostic and clinical implications.^3^ In patients with heart failure with reduced ejection fraction (HFrEF), a meta-analysis found the prevalence of RVD to be ∼48%, similar to the findings we present here. In contrast, the prevalence of RVD in patients with HFpEF has been less well established, with estimates ranging between ∼10-50%, based on the RV measure used.^6,16^ In our study, ∼30% of patients diagnosed with HFpEF had concomitant RVD. This difference likely reflects our more composite definition of RV dysfunction, compared to other studies’ reliance on a single echocardiographic variable. Interestingly, longitudinal assessment of patients with HFpEF who underwent serial echocardiography demonstrated a significant increase in the prevalence of isolated RVD and concomitant RVD-PH over time. These findings suggest that progressive right ventricular involvement may reflect ongoing adverse cardiopulmonary remodeling during HFpEF disease progression.^6^

Our work also highlights the operational characteristics of LLM-based platforms like ConceptExtract.AI that can add practical, real-world value across a variety of domains, even beyond clinical phenotyping (as we demonstrate here). End-to-end inference was completed at a median latency of 3.2 seconds per report at an approximate list-price cost of US$271 for the full cohort, a resource requirement that compares favorably to the time and personnel costs of equivalent manual chart review. The no-code design makes this capability accessible to clinical researchers and operational teams without informatics infrastructure, lowering the barrier to population-level phenotyping at scale. As we illustrate in Figure 2, potential applications are myriad. Cardiovascular imaging laboratories and their associated clinical enterprises generate terabytes of unstructured data daily, and a platform capable of continuously screening this output for any user-defined phenotypic pattern of interest (in our case, RVD with or without elevated pulmonary pressures, in the presence or absence of HF) could support a broad range of operational and clinical use cases. These include identification of patients who may benefit from specialist referral, prospective screening for clinical trial enrollment, generation of real-world evidence cohorts, administrative reporting, and quality improvement initiatives. The ability to define screening criteria in plain language rather than structured query logic makes such workflows adaptable in near realtime as clinical questions evolve, without requiring re-engineering of the underlying system. Together, these properties position ConceptExtract.AI as a scalable substrate for operationalizing complex clinical phenotypes from unstructured text across the full spectrum of research, clinical, and administrative applications.

Despite these promising results, there are limitations to this study. First, while this model robustly identifies echocardiographic signatures of RVD, it lacks validation against gold-standard invasive hemodynamic data. Similarly, conclusions based on the relationship between RVD and PH assume that echocardiographic features of PH correlate to a diagnosis based on invasive hemodynamics measurements, an assumption which may not always hold. Additionally, more than a quarter of TTE reports in our study lacked EF data, limiting our overall analysis of RV dysfunction in the presence of LV dysfunction. Future studies should focus on validating these echocardiographic phenotypes against a range of outcomes, including invasive hemodynamic data and, ultimately, clinically relevant, patient-centered endpoints. Of note, the operational cost and latency estimates reported here reflect GPT-4o list pricing at the time of analysis and a single institutional Azure deployment; these figures will vary across institutions, procurement agreements, and model versions.

## Conclusion

This study demonstrates that RVD and PH are common findings on echocardiography, identifiable both as isolated phenotypes and as clinically significant entities within the broader heart failure spectrum. These findings reinforce RVD, PH, and heart failure as interconnected but distinct entities that are not fully captured by any single echocardiographic variable and instead require a composite, contextual assessment to reliably identify. ConceptExtract.AI, a validated LLM-based extraction platform, enabled this phenotyping with higher fidelity than conventional rules-based approaches and can support population-level assessment at scale. The platform’s no-code design, low per-record cost, and real-time throughput extend its utility beyond research, enabling operational deployment for continuous clinical screening, trial enrollment, quality improvement, and real-world evidence generation. Further validation against complementary imaging, invasive hemodynamics, and clinical outcomes is needed to establish full clinical utility.

## Data Availability

All data produced in the present study are available upon reasonable request to the authors.

